# Automating Imaging Biomarker Analysis for Knee Osteoarthritis Using an Open-Source MRI-Based Deep Learning Pipeline

**DOI:** 10.1101/2025.02.21.25322094

**Authors:** Ananya Goyal, Francesca Belibi, Vyoma Sahani, Rune Pedersen, Yael Vainberg, Ashley Williams, Constance Chu, Bryan Haddock, Garry Gold, Akshay Chaudhari, Feliks Kogan, Anthony Gatti

## Abstract

Osteoarthritis (OA) is a leading cause of chronic disability worldwide, with knee OA being the most prevalent form. Quantitative assessment of knee joint tissues using Magnetic Resonance Imaging (MRI) and Positron Emission Tomography (PET) has the potential to enhance OA diagnosis and progression tracking. However, current methodologies for segmenting and extracting quantitative metrics from knee joint images are time-consuming, require extensive expertise, and suffer from inter- and intra-reader variability, limiting their clinical translation.

In this study, we present and validate a fully automated AI-based pipeline for comprehensive segmentation and quantitative analysis of knee joint tissues from MRI and PET images. Our pipeline segments key joint tissues, including femoral, tibial, and patellar bones, femoral, patellar, and tibial cartilage, and medial and lateral menisci, using a deep learning-based segmentation model. Furthermore, the pipeline enables automated extraction of critical quantitative OA biomarkers, including regional cartilage thickness and T2 relaxation times, meniscus volume, a neural shape model-derived OA bone shape score, and [^18^F]NaF PET-based measures of subchondral bone metabolism.

To evaluate the segmentation performance, we validated automated segmentations against manual segmentations from two annotators and compared derived quantitative metrics. Results demonstrated high segmentation accuracy, with DSC values ranging from 0.84 to 0.98 across different tissues. Automated quantitative measurements exhibited good to excellent agreement with manual-derived values for most metrics, with ICC values exceeding 0.89. Our findings suggest that the proposed AI-based pipeline provides a robust, open-source tool for efficient and reproducible quantitative analysis for knee OA studies, accelerating research and clinical adoption of whole-joint quantitative imaging.

## 1. INTRODUCTION

Osteoarthritis (OA) is the leading cause of chronic disabilities in the US and affects 595 million people worldwide [1]. OA is characterized by chronic pain and loss of mobility, both of which reduce productivity and quality of life at a tremendous cost to society. The knee is the joint most frequently impacted by OA, and knee OA cases are projected to increase by 75% between 2020 and 2050 [2].

OA is a whole-joint disease, involving the degeneration of multiple musculoskeletal tissues such as cartilage, meniscus, bone, synovium, and ligaments [3]. Magnetic Resonance Imaging (MRI) can provide quantitative morphological (structural) and compositional information. MRI-based information about articular cartilage thickness and quantitative T2 relaxation times have been shown to be predictive of OA disease progression [4-6]. Meniscal volume is representative of early meniscus degeneration, a known OA risk factor [7,8]. Shape model-based measures of femur bone shape can quantify osteophytes and predict knee pain [9-11]. Furthermore, [^18^F]NaF PET imaging can evaluate bone remodeling, with previous work showing elevated PET uptake in people with knee OA compared to healthy controls [12-13].

While all of these quantitative measures provide a more holistic approach to studying and predicting progression of OA, they are still challenging to use for research, and their clinical translation is limited. This is due to multiple factors. Manual or semi-automated tissue segmentation is affected by inter- and intra-user variability and is extremely time-consuming, taking hours for a single joint [14]. Even when segmentations are available, extraction of each of these metrics requires highly specialized skills and post-processing techniques such as fitting quantitative MRI parameter maps [15], image registration [16], surface extraction and geometry processing for cartilage thickness calculation [17], as well as an entire set of tools and methods for fitting shape models and computing bone-shape features [18]. Furthermore, most semi-automated or fully automated segmentation and analysis methods lack validation of quantitative measures and only compare results to a single manual annotator. To accelerate research and progress towards clinical adoption of whole joint quantitative MRI, we require robust open-source tools for fully automated segmentation and quantitative analysis of multiple knee tissues.

The purpose of this study was to develop and evaluate a fully automated AI-based pipeline for comprehensive MRI-based segmentation and quantitative analysis of multiple knee tissues from multi-modal MR and PET images. We comprehensively validate segmentations of the primary knee tissues (cartilage, bone, meniscus) and holistic quantitative metrics including regional cartilage thickness and T2 relaxation times, meniscus volume, a neural shape model-based OA-bone shape score, and [^18^F]PET-based measures of subchondral bone metabolism. We compared segmentation and clinical measures of two manual annotators with those of our automated pipeline, which is shared as a unified ‘KneePipeline’ https://github.com/gattia/KneePipeline) [19] and shown in Figure 1.

**Figure 1:**
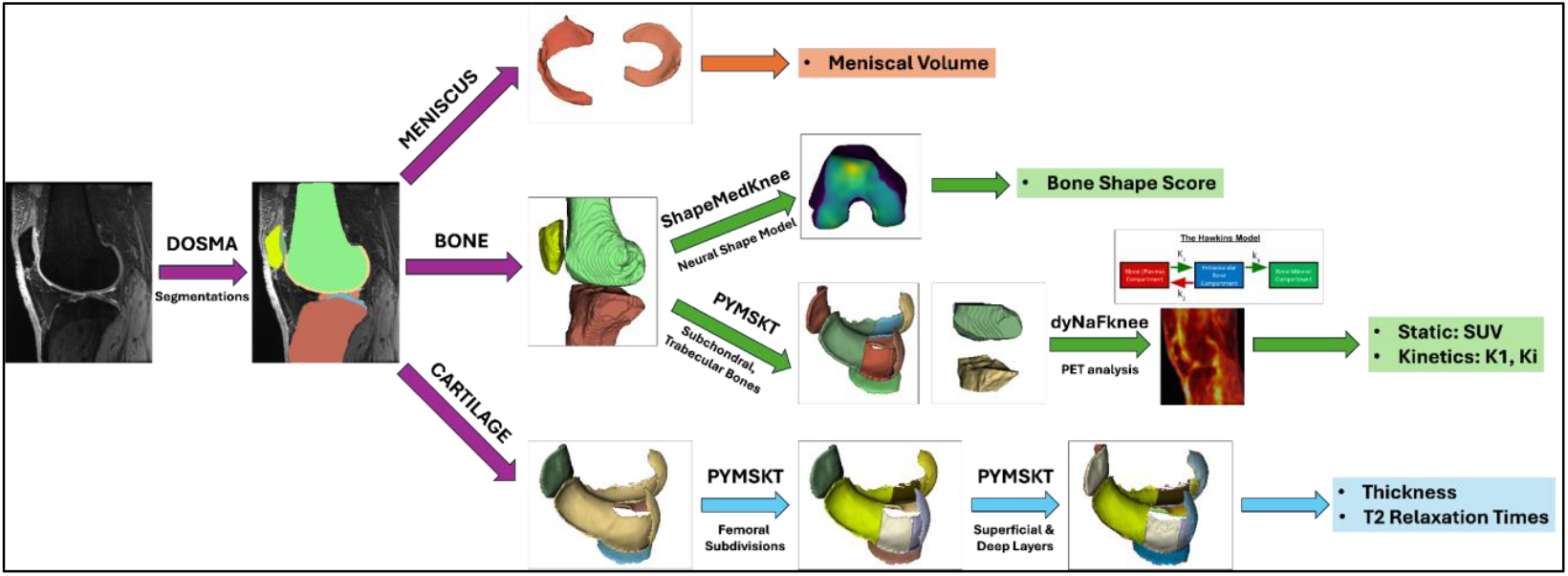
Overview of the KneePipeline [19]. Using a qDESS volume as input, DOSMA [17] automatically segments knee bones (femur, tibia, patella), cartilage (femoral, patellar, medial and lateral tibial), and menisci (lateral and medial). Each of the tissues has a sub-pipeline for quantitative processing. Overall, the pipeline allows for calculation of cartilage T2 and thickness, meniscus volumes, bone shape scores, and [^18^F]NaF PET SUV and kinetic measures.

## 2. METHODS

We curated a large dataset to train a robust deep learning model for automated segmentation and validated the segmentation performance using a prospectively acquired dataset of 20 image volumes. We compared automated segmentations with manual segmentations from two annotators in terms of segmentation quality and clinically relevant outcome measures. As part of this process, we developed a comprehensive segmentation and post-processing pipeline including bone, cartilage and meniscus analysis from MR and PET images, which we publicly share on GitHub [19] and via a 3D Slicer module.

### 2.1 Image datasets

Quantitative osteoarthritis outcomes are typically extracted from fat saturated gradient echo images [20, 21]. To train a robust segmentation model, we curated a training dataset with 176 standard double-echo steady state (DESS) images from Siemens 3T scanners at four sites [22], 155 quantitative DESS (qDESS) images from one of two 3T GE MR750 scanners [23], and 16 qDESS images from subjects with known anterior cruciate ligament reconstruction from a 3T Siemens Magnetom [8]. Validation was performed on qDESS images acquired from a prospective dataset from a 3T GE MR750 scanner with the following parameters: repetition time (TR) 18.24ms, echo times (TEs) 6.04ms and 30.44ms, matrix size 512×512, field of view 16cm, voxel size 0.3125×0.3125×1.5 mm^3^, 118 slices per TE. qDESS produces two echoes (S+ and S-), where S+ has high SNR with T1/T2 weighting while S-has fluid sensitivity with higher T2 weighting. Analytical signal equations fit to the two echoes can be used to create cartilage T2 relaxation time maps [20]. All participants provided written consent for the study.

### 2.2 Neural network for automated segmentation

We trained a 2D convolutional neural network to segment three bones (femur, tibia, patella), four cartilage volumes (femoral, medial and lateral tibial, and patellar), and two meniscal volumes (medial and lateral). The network used a 2D U-Net style architecture previously described [24] with an image input size of 512×512, deep supervision, and an output of 512×512 with 9 feature channels for the 9 tissues. The network was trained using data augmentation, including random in-plane rotations within 6° and translations within ±20%, batch normalization, and dropout of 0.2. A loss comprised of the negative of the sum of individual tissue dice similarity coefficients (DSC) was optimized using the Adam optimizer [25], a batch size of 12, a learning rate of 1e-4.5, and early stopping when the loss improved <0.001 over 10 epochs. The network was implemented and trained using Keras 2 [26].

### 2.3 Division of segmentations into subregions

#### Cartilage

Femoral cartilage was further subdivided into five subregions (anterior, medial and lateral central, medial and lateral posterior) using the Python library, *PYMSKT* [27] for cartilage thickness and T2 relaxation time analysis. This subregion division was based on established region definitions and anatomical landmarks used for osteoarthritis clinical trials [28]. Further, all cartilage regions were separated into deep and superficial layers by computing the relative depth of each voxel, with a depth of 0.0 being on the bone surface and 1.0 being on the articular surface; depth <0.5 was classified as deep and >=0.5 as superficial. In total, there were eight cartilage subregions, and each region was further differentiated into superficial and deep layers.

#### Bone

From the whole bone segmentations, sub-regional masks of the subchondral and trabecular bones were created for [^18^F]PET analysis.

<u>Subchondral bones</u> (femur, tibia, patella) were created by selecting the bone voxels that lay within a 3mm thick threshold of the cartilage surface. This thickness was chosen to account for the low resolution and signal spilling out in PET images, which is 1.3×1.3×2.78mm^3^.

Subchondral bone regions were sub-divided into anatomical regions of interest using the *pymskt* cartilage subdivisions as defining regions: anterior, medial and lateral central, medial and lateral posterior for the femur, medial and lateral tibial, and patellar, resulting in eight total regions.

<u>Trabecular bones</u> (femur, tibia) subregion was created by labeling all bone voxels greater than 9mm from the bone surface.

### 2.4 Clinical quantitative measurements

Once images are segmented, they require considerable post-processing to extract quantitative measures of joint health. Below, we describe the methods used to extract quantitative measures automatically. All described methods are freely shared on GitHub [19], enabling comprehensive knee tissue analysis.

#### Cartilage

1) T2 Relaxation Times

Cartilage T2 relaxation times were computed for each voxel using a previously defined analytical approach based on the qDESS MRI acquisition [17,20]. Mean T2 relaxation times were computed voxel-wise and then averaged over the eight segmentation-defined regions of interest. For each region, T2 relaxation times were calculated for whole cartilage, as well as deep and superficial layers.

2) Mean Thickness

Cartilage thickness was computed by converting segmentation masks into surface meshes and computing the overlying articular cartilage surface distance normal to the bone surface. Concurrently, each vertex was labelled by the overlay cartilage subregion (e.g., medial central femur, or lateral tibia). Average cartilage thickness was computed as the mean thickness for all vertices with a given label.

#### Bone

1) BScore

A BScore is a shape model-derived measure of whether a bone has OA-like features, with a higher score indicating the bone is more characteristic of OA [11]. We previously developed a neural shape model that encodes combined femoral bone and cartilage shape and trained it on 6,325 knees from the Osteoarthritis Initiative. We then created a neural shape model-based metric, BScore, using a previously established methodology [11]. We fit this neural shape model to each subject’s femoral bone and cartilage surfaces and computed their BScore [18]. The neural shape model is open-source [29] and is implemented using PyTorch [30].

2) Subchondral Bone Metabolism from [^18^F]NaF PET Imaging

Subregional bone masks (subchondral and trabecular bones) derived from whole bone segmentations were used to calculate [^18^F]NaF PET mean and maximum standardized uptake values (SUV) and bone perfusion (K1) and total bone mineralization (KiNLR) derived from the Hawkin’s three compartment model [31] of [^18^F]NaF PET skeletal kinetics. The PET image dataset & processing pipeline are described in the Supplementary Text 1.

#### Meniscus

1) Volume

Meniscus volume was calculated for both medial and lateral menisci as the product of the sum of total voxels in the segmentation and voxel volume.

### 2.5 Validation of automated segmentations

#### Comparison study with Manual Segmentations

To validate the automated bone segmentations from DOSMA, two annotators with one (F.B.) and two years (V.S.) of experience performed manual segmentations for 20 subjects (10 with self-reported knee OA and 10 controls). Unilateral qDESS knee scans (Section 2.1) were used for segmentation. After segmentation, these regions were subdivided into further bone and cartilage sub-regions as described earlier (Section 2.3), and the quantitative measures were calculated. Lastly, to test the performance of the automated segmentations in the epiphyseal region, the whole bone segmentations were sectioned to exclude the long bone for the tibia and femur.

#### Comparison of segmentation quality

To test the accuracy of our automated segmentations, three segmentation metrics were calculated for each of the segmented regions and sub-regions: Dice similarity coefficient (DSC), average symmetric surface distance (ASSD), and volume difference (VD), using NumPy and SimpleITK in Python [32,33].

#### Comparison of quantitative metrics

To evaluate the downstream effects of manual versus automated segmentation, we compared the quantitative metrics described previously. Intraclass correlation coefficients (ICC) and normalized root mean squared error (NRMSE) over the mean were calculated to evaluate the agreement between manual and automated measurements. Measures were interpreted as poor agreement for ICC <0.5, moderate agreement for ICC 0.5-0.75, good agreement for ICC 0.75-0.9, and excellent for ICC>0.9.

## 3. RESULTS

### 3.1 Patient Demographics

The validation study included 10 subjects with self-reported, symptomatic knee OA (5 male, 5 female, average age 44±17 years) and 10 asymptomatic control subjects (5 male, 5 female, average age 54±7 years).

### 3.2 Automated Segmentation Performance

Image metrics (DSC, ASSD, VD) for segmentation performance between the two annotators’ manual segmentations and automated segmentations are shown in Table 1 and Figure 2, for cartilage, bones and menisci. The performance of femoral cartilage subdivisions and trabecular bones are included in Supplementary Table S1.

**Table 1.** Comparison of segmentation performance between manual ground truth segmentations (Reader 1-R1, Reader 2-R2) and automated segmentations (Au). Data are mean (standard deviation), and the best performing out of all three are bolded. ASSD: Average symmetric surfaces distance; DSC: Dice similarity coefficient.

| Region | DSC |  |  | ASSD (mm) |  |  | Volume Difference (%) |  |  |
| --- | --- | --- | --- | --- | --- | --- | --- | --- | --- |
|  | R1 vs Au | R2 vs Au | R1 vs R2 | R1 vs Au | R2 vs Au | R1 vs R2 | R1 vs Au | R2 vs Au | R1 vs R2 |
| **Whole Bones** |  |  |  |  |  |  |  |  |  |
| Femur | 0.98<br>(0.01) | 0.97<br>(0.01) | **0.98**<br>**(0.01)** | 0.26<br>(0.15) | 0.32<br>(0.15) | **0.23**<br>**(0.08)** | 1.97<br>(1.03) | 3.26<br>(2.06) | **-1.36**<br>**(1.62)** |
| Tibia | **0.98**<br>**(<0.01)** | 0.97<br>(<0.01) | 0.98<br>(<0.01) | **0.19**<br>**(0.03)** | 0.22<br>(0.05) | 0.19<br>(0.04) | 1.95<br>(1.05) | 3.67<br>(0.87) | **-1.78**<br>**(1.30)** |
| Patella | 0.95<br>(0.01) | 0.95<br>(0.01) | **0.96**<br>**(0.01)** | 0.25<br>(0.09) | 0.25<br>(0.06) | **0.18**<br>**(0.05)** | 8.00<br>(2.72) | 9.58<br>(2.26) | **-1.77**<br>**(2.87)** |
| Femur<br>(Epiphyseal Region) | **0.98**<br>**(<0.01)** | 0.98<br>(<0.01) | 0.98<br>(<0.01) | 0.15<br>(0.05) | 0.19<br>(0.06) | **0.15**<br>**(0.05)** | **1.30**<br>**(0.67)** | 2.57<br>(1.34) | -1.32<br>(1.25) |
| Tibia<br>(Epiphyseal Region) | **0.98**<br>**(<0.01)** | 0.98<br>(<0.01) | 0.98<br>(<0.01) | **0.13**<br>**(0.03)** | 0.16<br>(0.04) | 0.14<br>(0.05) | **1.60**<br>**(0.75)** | 3.19<br>(0.79) | -1.65<br>(1.16) |
| **Whole Cartilage** |  |  |  |  |  |  |  |  |  |
| Femoral | **0.89**<br>**(0.02)** | 0.87<br>(0.02) | 0.86<br>(0.02) | **0.13**<br>**(0.05)** | 0.16<br>(0.05) | 0.17<br>(0.05) | -6.22<br>(6.03) | -6.62<br>(6.45) | **0.09**<br>**(7.47)** |
| Medial Tibial | **0.87**<br>**(0.02)** | 0.86<br>(0.03) | 0.84<br>(0.03) | **0.13**<br>**(0.05)** | 0.17<br>(0.08) | 0.18<br>(0.06) | -9.10<br>(7.44) | -9.63<br>(5.18) | **0.34**<br>**(7.27)** |
| Lateral Tibial | **0.89**<br>**(0.03)** | 0.87<br>(0.03) | 0.87<br>(0.03) | **0.17**<br>**(0.09)** | 0.21<br>(0.09) | 0.21<br>(0.07) | -5.90<br>(6.42) | -8.37<br>(7.71) | **1.99**<br>**(6.62)** |
| Patellar | **0.91**<br>**(0.02)** | 0.88<br>(0.03) | 0.89<br>(0.02) | **0.11**<br>**(0.06)** | 0.16<br>(0.09) | 0.16<br>(0.08) | -5.80<br>(6.04) | -8.23<br>(4.91) | **2.11**<br>**(6.27)** |
| **Meniscus** |  |  |  |  |  |  |  |  |  |
| Medial | **0.88**<br>**(0.02)** | 0.88<br>(0.03) | 0.85<br>(0.03) | **0.20**<br>**(0.09)** | 0.21<br>(0.09) | 0.26<br>(0.08) | -3.71<br>(4.50) | **-1.64**<br>**(5.95)** | -2.37<br>(7.57) |
| Lateral | **0.89**<br>**(0.02)** | 0.87<br>(0.04) | 0.85<br>(0.03) | **0.17**<br>**(0.05)** | 0.27<br>(0.23) | 0.30<br>(0.22) | -8.62<br>(3.86) | -10.73<br>(5.49) | **1.71**<br>**(5.36)** |
| **Subchondral Bones** |  |  |  |  |  |  |  |  |  |
| Anterior Femoral | **0.90**<br>**(0.02)** | 0.88<br>(0.02) | 0.89<br>(0.02) | **0.18**<br>**(0.07)** | 0.21<br>(0.08) | 0.20<br>(0.07) | **-0.20**<br>**(6.88)** | -1.52<br>(5.60) | 1.22<br>(5.68) |
| Medial Central Femoral | **0.87**<br>**(0.03)** | 0.87<br>(0.03) | 0.86<br>(0.04) | 0.21<br>(0.10) | **0.20**<br>**(0.07)** | 0.21<br>(0.12) | -0.87<br>(6.02) | -0.61<br>(5.34) | **-0.36**<br>**(5.25)** |
| Lateral Central Femoral | 0.89<br>(0.04) | **0.89**<br>**(0.03)** | 0.87<br>(0.03) | 0.19<br>(0.13) | **0.16**<br>**(0.07)** | 0.20<br>(0.09) | **-0.30**<br>**(2.85)** | 1.42<br>(4.04) | -1.90<br>(5.02) |
| Medial Posterior Femoral | **0.91**<br>**(0.02)** | 0.88<br>(0.02) | 0.88<br>(0.03) | **0.18**<br>**(0.12)** | 0.22<br>(0.14) | 0.21<br>(0.10) | **-1.31**<br>**(5.22)** | 1.60<br>(4.89) | -3.05<br>(4.61) |
| Lateral Posterior Femoral | **0.90**<br>**(0.03)** | 0.88<br>(0.03) | 0.87<br>(0.03) | **0.16**<br>**(0.08)** | 0.20<br>(0.09) | 0.21<br>(0.09) | -2.48<br>(6.47) | **0.07**<br>**(4.60)** | -2.66<br>(6.51) |
| Medial Tibial | **0.90**<br>**(0.02)** | 0.88<br>(0.02) | 0.88<br>(0.02) | **0.21**<br>**(0.15)** | 0.23<br>(0.09) | 0.26<br>(0.14) | 0.13<br>(3.65) | **-0.12**<br>**(4.62)** | 0.13<br>(4.10) |
| **Lateral Tibial** | **0.90**<br>**(0.03)** | 0.87<br>(0.02) | 0.88<br>(0.03) | **0.21**<br>**(0.12)** | 0.28<br>(0.11) | 0.25<br>(0.10) | 1.10<br>(5.31) | **-0.74**<br>**(6.20)** | 1.60<br>(6.21) |
| **Patellar** | **0.92**<br>**(0.01)** | 0.88<br>(0.03) | 0.89<br>(0.03) | **0.09**<br>**(0.03)** | 0.16<br>(0.06) | 0.14<br>(0.06) | 2.28<br>(2.19) | 3.17<br>(2.35) | **-0.94**<br>**(2.23)** |

**Figure 2.**
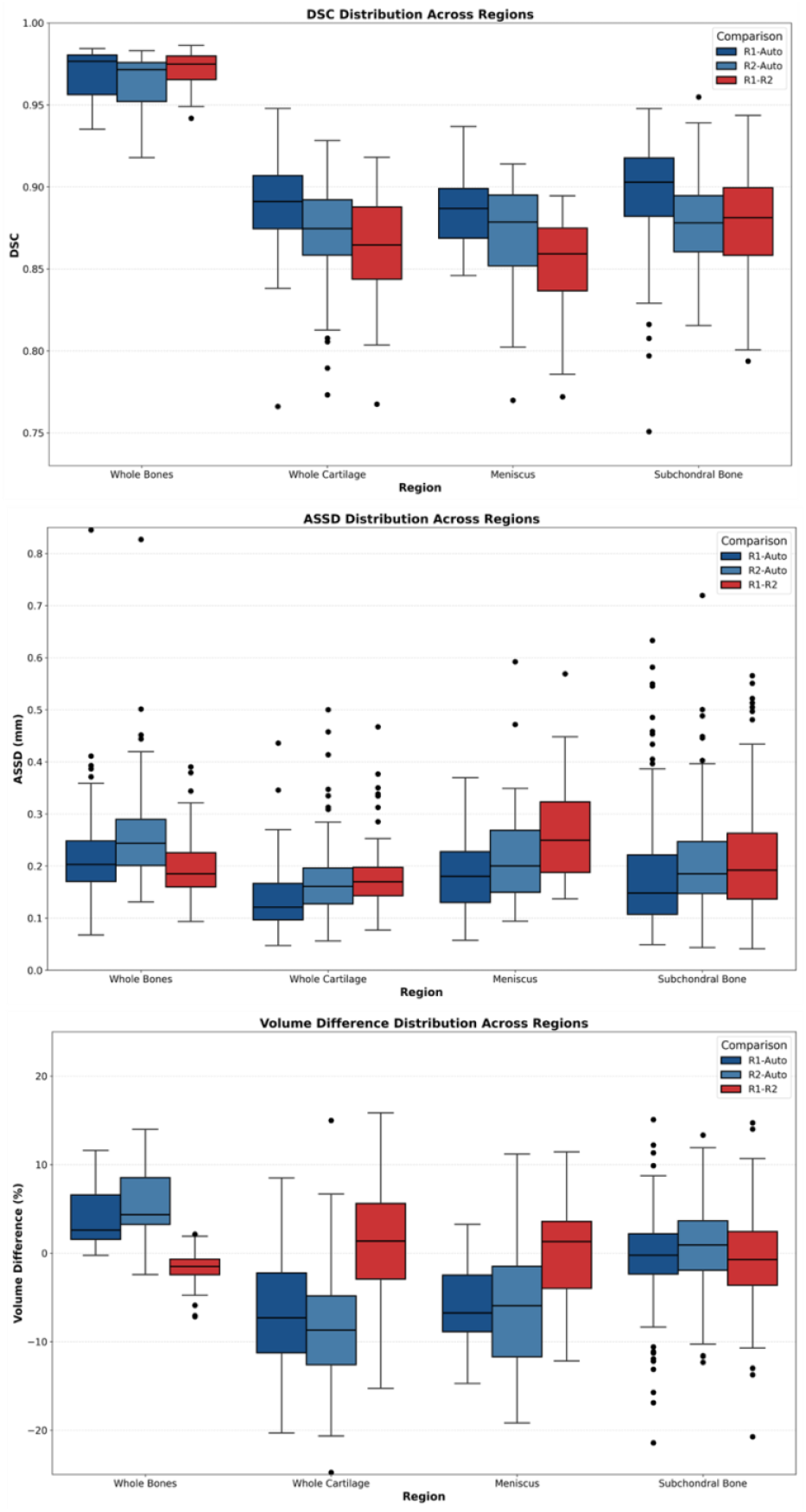
Values of DSC, ASSD, and volume differences across subjects and readers for different groups of segmentations. Blue boxplots are manual versus automated, and red are inter-reader comparisons.

#### Whole bones

The whole bone regions (femur, tibia, patella) show consistently high DSC (0.95 to 0.98), indicating excellent accuracy across all reader comparisons. The ASSD values are low (0.13 to 0.32 mm), suggesting minimal deviation in surface distance between segmented bones. Volume differences range from small negative to moderate positive values (1.30% to 9.58%), with the patella showing the highest volume difference.

#### Cartilage

Cartilage regions show lower DSC values (0.84 to 0.91) than bone regions. Yet, the ASSD values are smaller than in bone (0.11 to 0.21 mm). The volume differences are often negative between manual and automated segmentations, reflecting a tendency for the automated segmentations to underestimate volume compared to the manual segmentations.

#### Subchondral bones

Subchondral bone regions exhibit high DSC (0.86 to 0.91) and small ASSD (0.14 to 0.28 mm) values, indicating good agreement, with the lateral tibial subchondral bone showing the highest ASSD. Volume differences were generally small to moderate, ranging from -3.05% to 3.17%.

#### Menisci

Meniscus regions exhibit DSC values in the range of 0.85 to 0.89 and ASSD of 0.17 to 0.30 mm, which is less than in-plane resolution. Volume differences show larger variation, with regional averages for the lateral meniscus ranging from -10.73% to 1.71% and for the medial meniscus ranging from -3.71% to -1.64%.

### 3.3 Quantitative Metrics

To evaluate downstream performance of the manual versus automated segmentations, quantitative metrics for each tissue were calculated (Supplementary Table S2). Agreement between raters for selected results are shown in Table 2 and Figure 3 (full results are provided in Supplementary Table S3).

**Table 2.** Results of Intra-class correlation coefficient (ICC) analysis and normalized root mean squared error (NRMSE) (normalized over the range) analysis for quantitative metrics for cartilage, bone, and menisci, across the three segmentation groups. Only some regions are included here, rest are in Supplementary Table S3.

| Region | ICC |  |  | Normalized RMSE |  |  |
| --- | --- | --- | --- | --- | --- | --- |
|  | R1 vs Auto | R2 vs Auto | R1 vs R2 | R1 vs Auto | R2 vs Auto | R1 vs R2 |
| **Bscore** |  |  |  |  |  |  |
| Femur | 0.92 | 0.46 | 0.49 | 0.15 | 0.41 | 0.38 |
| **Meniscal Volume (mm3)** |  |  |  |  |  |  |
| Medial Meniscus | 0.97 | 0.97 | 0.97 | 0.06 | 0.07 | 0.07 |
| Lateral Meniscus | 0.93 | 0.90 | 0.98 | 0.09 | 0.11 | 0.05 |
| **Cartilage Thickness (mm)** |  |  |  |  |  |  |
| Medial Central Femoral | 0.88 | 0.81 | 0.83 | 0.07 | 0.10 | 0.09 |
| Patellar | 0.91 | 0.87 | 0.93 | 0.07 | 0.08 | 0.06 |
| **Whole Cartilage T2 Relaxation Times (ms)** |  |  |  |  |  |  |
| Medial Central Femoral | 0.99 | 0.96 | 0.96 | 0.02 | 0.03 | 0.03 |
| Patellar | 0.98 | 0.90 | 0.94 | 0.02 | 0.04 | 0.03 |
| **SUVmean** |  |  |  |  |  |  |
| Medial Central Femoral | 1.00 | 1.00 | 1.00 | 0.04 | 0.05 | 0.06 |
| Patellar | 1.00 | 1.00 | 1.00 | 0.01 | 0.02 | 0.01 |
| **KiNLR** |  |  |  |  |  |  |
| Medial Central Femoral | 0.99 | 0.99 | 1.00 | 0.08 | 0.09 | 0.05 |
| Patellar | 1.00 | 1.00 | 1.00 | 0.04 | 0.06 | 0.06 |

**Figure 3.**
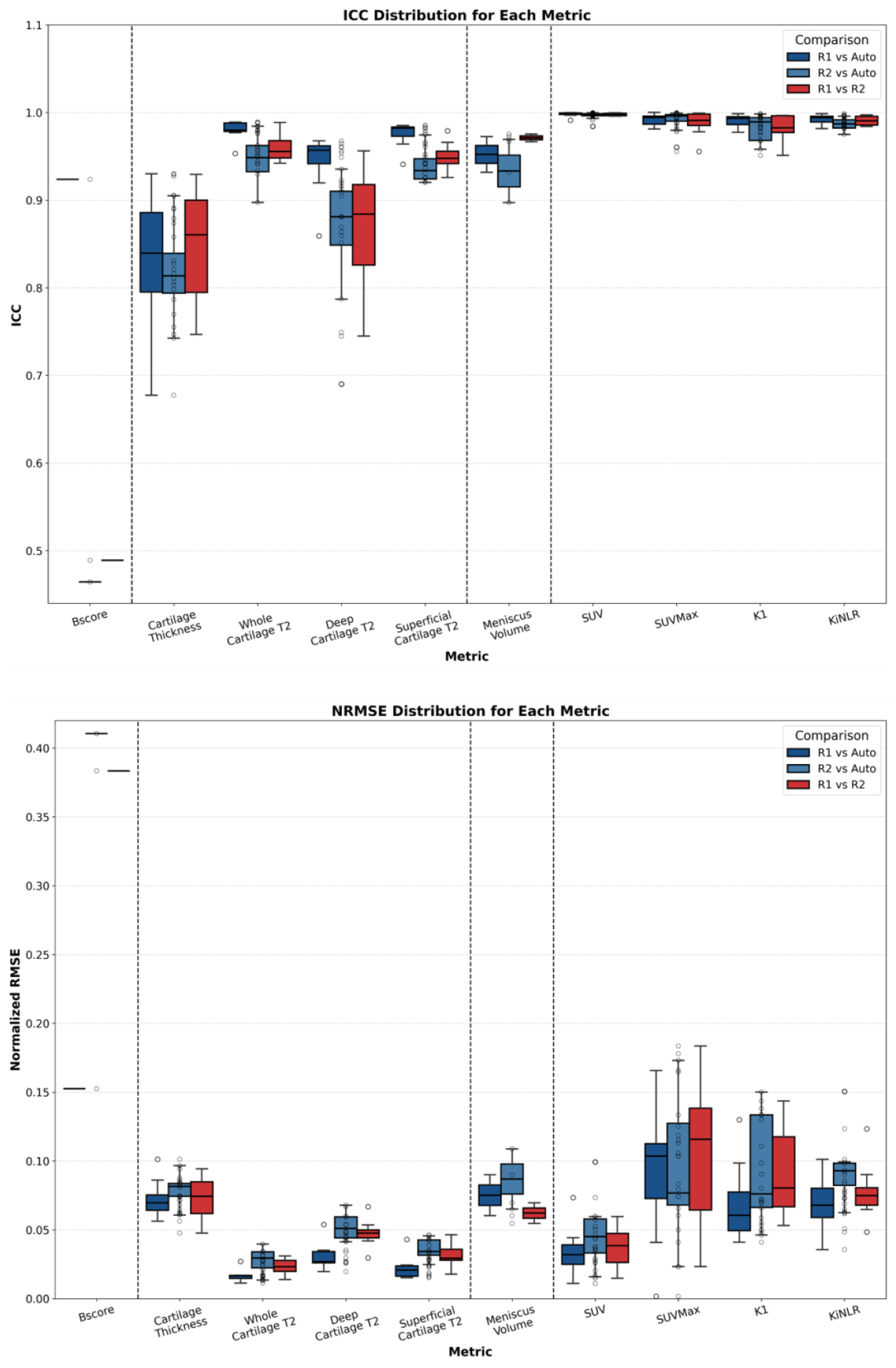
Average Intra-class correlation coefficient (ICC) and normalized root mean squared error (NRMSE) across subregions for quantitative metrics of cartilage, bone, and menisci.

#### Cartilage

1) T2 Relaxation Times

The whole cartilage T2 values showed ICC values between 0.95 to 0.99 (Reader 1) and 0.89 to 0.98 (Reader 2) for readers versus automated, and 0.94 to 0.98 for inter-reader comparisons. Similarly, superficial cartilage had excellent agreement between readers ranging between 0.92 to 0.98, while for deep cartilage, there is more variability in T2, ranging from 0.69 to 0.96. The lower values were for Reader 2 versus automated and Reader 1 versus Reader 2. The NRMSE for whole cartilage T2 ranged from 0.01 to 0.04, showing the smallest errors for Reader 1 versus automated, followed by Reader 1 versus Reader 2 and Reader 2 versus automated, which is consistent for deep and superficial T2 as well. Sample images showing segmented cartilage and T2 maps are in Figure 4.

**Figure 4.**
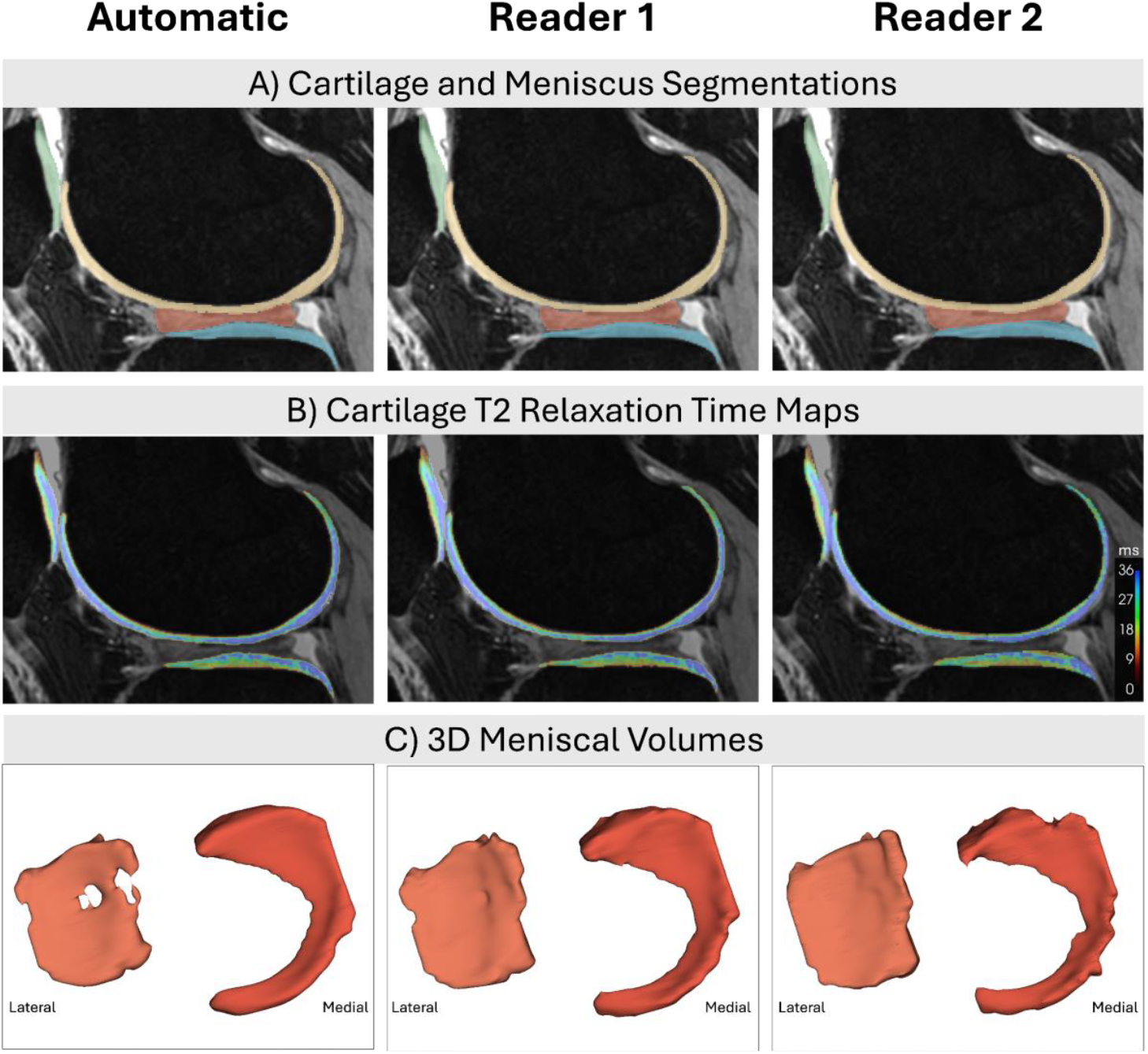
Sample cartilage (femoral, patellar, and lateral tibia) and lateral meniscus segmentations are shown for a subject in their sixties with self-reported knee osteoarthritis. Corresponding cartilage T2 relaxation time maps are shown in panel B. Of note, in panel C, are the medial and lateral meniscus volumes. This volunteer has a discoid lateral meniscus, which is an abnormally shaped meniscus that is more likely to be injured than a typical C-shaped meniscus. The automated segmentation for the lateral meniscus shows some missing areas, which is not seen in the manual segmentations.

2) Mean Thickness

Mean cartilage thickness ICC values per region of interest are comparable for three pairs of segmentations, ranging from 0.68 to 0.93 across the groups, while NRMSE was also comparable across the pairs (range 0.06 to 0.10).

#### Meniscus

1) Volume

For both medial and lateral meniscus volumes, the measurements between Reader 1 and Reader 2 as well as Reader 2 and automated segmentations show excellent agreement, with low NRMSE (ICC: 0.93 to 0.97, NRMSE: 0.05 to 0.1). The lateral meniscus, however, shows a good agreement (ICC of 0.90 for Reader 2 versus automated). Sample images showing segmented menisci and volumes are in Figure 4.

#### Bone

1) BScore

The Reader 1-automated measurements show very excellent agreement (ICC:0.92, NRMSE:0.15), while both the Reader 1 versus Reader 2 and Reader 2 versus automated comparisons exhibit poor agreement (ICC: 0.49 and 0.46 respectively) with relatively higher error (NRMSE:0.38 and 0.41), particularly between Reader 2 and automated. Figure 5 shows variations in bone and cartilage shape.

**Figure 5.**
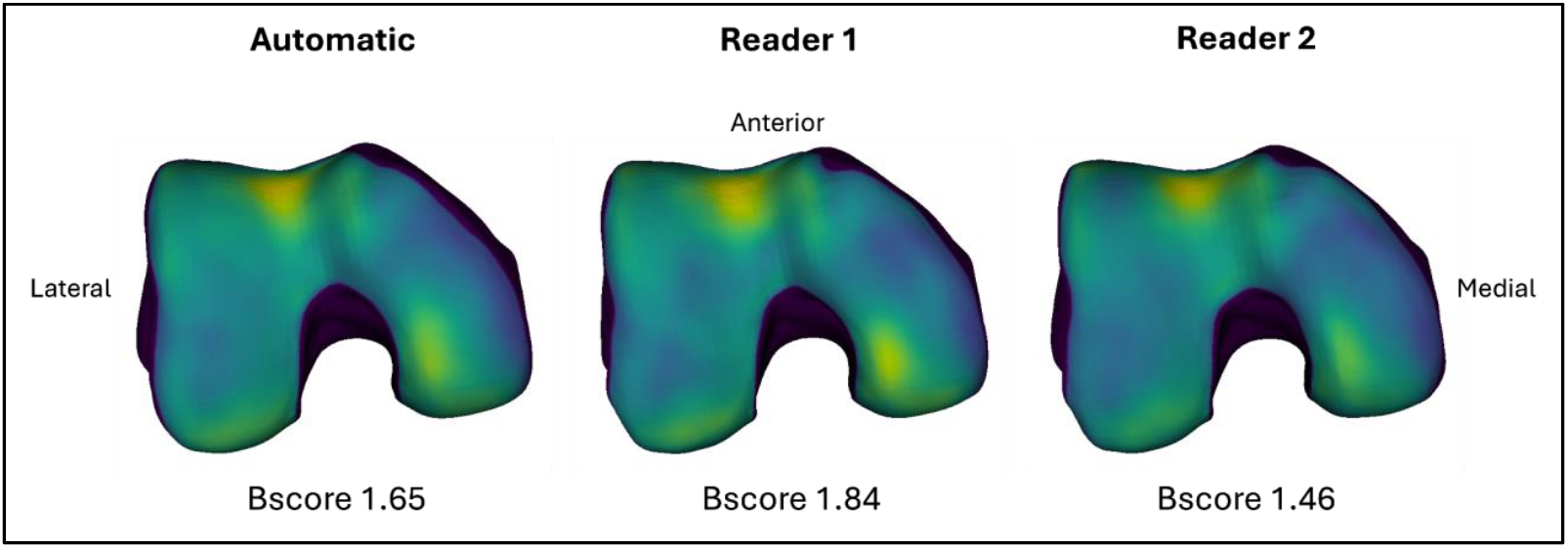
Bone shape models for one subject are shown below, for all three segmentations. The colormap represents projection of cartilage thickness onto the bone surface, brighter color indicates thicker cartilage. The BScores for this subject range from 1.46 to 1.84. A mean BScore of 0 is equivalent to the average healthy bone from the training dataset, with 1 unit equivalent to 1 standard deviation in the healthy group. This subject is thus on the upper end of the healthy spectrum being ∼1.65 standard deviations from the mean. This matches the observed shape and cartilage thickness, which shows a broad bone surface consistent with early osteoarthritis, and the beginning of cartilage thinning on the medial side.

2) Subchondral Bone SUV and SUVmax

The agreement between all three comparison groups is between 0.98 to 1, with relatively low NRMSE (0.01 to 0.09) across both values.

3) Subchondral Bone K1 and KiNLR

For both kinetic measures, all three comparisons show very good to excellent agreement across most measurements. The lowest agreement and highest errors are seen in the medial central femoral subchondral bone, but these still maintain strong agreement (ICC: 0.96 to 0.97, NRMSE: 0.13 to 0.15). Patella, tibia, and femur measurements generally exhibit excellent consistency (ICC: 0.96 to 0.99) with low errors (NRMSE: 0.03 to 0.15).

## 4. DISCUSSION

In this study, we showed that our fully automated AI-based pipeline can comprehensively segment and accurately perform quantitative analysis of multiple knee tissues from MRI and PET images. We are the first to compare deep learning segmentations with manual segmentations from two annotators and to show that automated segmentations are on par, if not better, than inter-reader segmentations.

Furthermore, we evaluated the downstream effects of these automated segmentations on all widely studied quantitative knee metrics such as cartilage T2 and thickness, bone shape, meniscus volume, and PET SUV and kinetic measures. We found that the majority of the metrics derived from automated segmentations are comparable to those derived from manual segmentations, as shown by good to excellent reliability and high accuracy. The best segmentation performance (DSC, ASSD, volume difference) and quantitative outcome measures (ICC, NRMSE) were commonly between the automated segmentation and one of the raters. Overall, our standardized open-source AI-based pipeline provides an easy way to perform automated whole joint analysis in minutes, as compared to the hours it typically takes to complete.

The segmentation accuracy of the model, measured by DSC, ASSD and volume differences, was comparable to the manual segmentations performed by two annotators. Our results align with previous studies that have developed automated segmentation pipelines for knee MRI, focusing on either both bone and cartilage or singular bone, cartilage, or meniscus segmentations. Prior research using deep learning-based segmentation has reported DSC scores in the ranges of 0.96 to 0.99 for knee bones [34-36], 0.79 to 0.89 for knee cartilage [34-36] and 0.66 to 0.89 for meniscus [37-39]. The vast majority of our DSC values fall within the upper range, further validating the quality of our segmentation approach.

### Cartilage metrics comparison

We found moderate to excellent agreement between manual and automated segmentations in calculating cartilage T2 relaxation times and thickness measures, matching findings of previous work in the field [40-42]. The automated-segmentation-derived cartilage regions showed T2 values highly correlated with those from manual segmentations, indicating that the automated segmentations capture the same differences between subjects as manual segmentations. T2 and thickness are the cornerstones for assessing early changes in cartilage in research and clinical trials [43]. The ability of our open-source, automated segmentation pipeline to compute cartilage thickness and T2 with negligible errors (NRMSE thickness <0.1, NRMSE T2 <0.04) has the potential to greatly reduce costs and increase sample sizes of osteoarthritis research and clinical trials, accelerating the field.

### Meniscus volume comparison

The meniscus volume obtained from automated segmentations was comparable to that obtained by the manual segmentations, with minimal errors and excellent agreement. Furthermore, meniscus volumes were comparable to those found in previous work using automated segmentation [37,39]. Given the role of meniscus morphology in knee biomechanics and osteoarthritis progression, accurate segmentation is crucial. While there are some minor differences in terms of volume differences of the lateral meniscus, overall, there was better segmentation DSC and ASSD agreement between the automated and each of the manual segmentations than between the two manual segmentations.

### Bone shape

The neural shape model-based BScores showed variable agreement between automated and manually segmented bones, ranging from poor to excellent agreement. BScores have been proposed as a more sensitive measure of osteoarthritis severity than X-ray-based Kellgren-Lawrence grades [11,44]. However, BScores have not widely been adopted as they require segmentation, shape modeling, and machine learning to determine and quantify this singular metric of osteoarthritis bone shape. Our open-source, automated, segmentation model, neural shape model, and BScore enable widespread use of shape analysis to quantify osteoarthritis severity. We hope the community can leverage this method to better understand the role of bone shape in osteoarthritis initiation and progression [45].

### Positron Emission Tomography (PET)

[^18^F]NaF PET-MRI has been of increasing interest for understanding osteoarthritis pathophysiology, showing promise in detecting subchondral bone remodeling and cartilage-bone interactions that contribute to OA disease progression [12,13,46,47]. However, calculation of PET metrics has yet to be standardized. In this work, we developed the first method for generating subchondral and trabecular bone subregions automatically, which we have open-sourced in our existing library, *PYMSKT* [27]. This is a crucial step for enabling reproducible knee PET-MRI research. Using these subchondral bone subregions obtained from only bone and cartilage segmentations, we found that SUV and kinetic measures of bone perfusion (K1) and total bone mineralization (KiNLR) extracted from automated segmentations were highly consistent with those from manual segmentations. An automated pipeline for PET analysis will be necessary to translate to bigger sample sizes. Furthermore, with the expanding use of other PET tracers such as [^18^F]-FDG and [^18^F]-FEPPA [48] for musculoskeletal analysis, our pipeline can be easily adapted for different PET applications. This is a critical step towards widespread use of quantitative PET-MRI for research, clinical trials, and potential clinical applications.

Our study had some limitations. We used a small validation dataset (20 knees) for evaluating performance of manual versus automated segmentations. However, we have two sets of manual segmentations per knee, and include segmentations of all major knee tissues, which provide more data than any prior work. Furthermore, our training set includes more than 300 knee MRIs from multiple vendors and sites. Additionally, our model was optimized for sagittal gradient echo images with fat saturation, and its performance on other MRI sequences remains to be explored. While the results suggest strong segmentation accuracy for most metrics, performance could be affected by variations in image resolution, contrast, and artifact presence. Future studies could extend this work by incorporating transfer learning to adapt the model for different MRI sequences and by incorporating other knee tissues such as tendons, ligaments, and fat tissue.

In this study, we developed and validated a fully automated open-source AI-based segmentation and analysis pipeline for MR and PET knee imaging, demonstrating its ability to accurately segment multiple knee tissues and perform comprehensive quantitative analysis with high accuracy. Compared to manual segmentation methods, our pipeline enables rapid, standardized analysis within minutes, making it well-suited for large-scale studies and clinical applications. Overall, this pipeline represents a significant step toward automating quantitative knee image analysis, thereby helping to make musculoskeletal imaging research more efficient and translatable.

## 5. CONCLUSION

Our open-source, AI-based, segmentation pipeline provides a fast, accurate, and reliable method for analyzing knee tissues from MRI and PET data. By automating segmentation and quantitative analysis, this approach has the potential to standardize knee MRI evaluation, reduce observer variability, and facilitate large-scale studies on osteoarthritis and joint health.

## Data Availability

All data produced in the present study are available upon reasonable request to the authors

https://github.com/gattia/KneePipeline

## REFERENCES

[1] GBD 2021 Osteoarthritis Collaborators. “Global, regional, and national burden of osteoarthritis, 1990-2020 and projections to 2050: a systematic analysis for the Global Burden of Disease Study 2021.” The Lancet. Rheumatology vol. 5,9 e508-e522. 21 Aug. 2023, doi:10.1016/S2665-9913(23)00163-7.

[2] Hawker, G. A., & King, L. K. (2022). The Burden of Osteoarthritis in Older Adults. Clinics in geriatric medicine, 38(2), 181–192. 10.1016/j.cger.2021.11.005.

[3] Poole A. R. (2012). Osteoarthritis as a whole joint disease. HSS journal : the musculoskeletal journal of Hospital for Special Surgery, 8(1), 4–6. 10.1007/s11420-011-9248-6.

[4] Eckstein, F et al. “Brief Report: Cartilage Thickness Change as an Imaging Biomarker of Knee Osteoarthritis Progression: Data From the Foundation for the National Institutes of Health Osteoarthritis Biomarkers Consortium.” Arthritis & rheumatology (Hoboken, N.J.) vol. 67,12 (2015): 3184–9. doi:10.1002/art.39324.

[5] Liebl, Hans et al. “Early T2 changes predict onset of radiographic knee osteoarthritis: data from the osteoarthritis initiative.” Annals of the rheumatic diseases vol. 74,7 (2015): 1353–9. doi:10.1136/annrheumdis-2013-204157.

[6] Prasad, A P et al. “T1ρ and T2 relaxation times predict progression of knee osteoarthritis.” Osteoarthritis and cartilage vol. 21,1 (2013): 69–76. doi:10.1016/j.joca.2012.09.011.

[7] Xu, Dawei et al. “Association between meniscal volume and development of knee osteoarthritis.” Rheumatology (Oxford, England) vol. 60,3 (2021): 1392–1399. doi:10.1093/rheumatology/keaa522

[8] Xu, Dawei et al. “Factors associated with meniscus volume in knees free of degenerative features.” Osteoarthritis and cartilage vol. 31,12 (2023): 1644–1649. doi:10.1016/j.joca.2023.08.003.

[9] Gatti, Anthony A et al. “Investigating acute changes in osteoarthritic cartilage by integrating biomechanics and statistical shape models of bone: data from the osteoarthritis initiative.” Magma (New York, N.Y.) vol. 35,5 (2022): 861–873. doi:10.1007/s10334-022-01004-8

[10] Gatti, Anthony A et al. “Predicting Chronic Knee Pain Using An Automated Mri-Based Bone And Cartilage Statistical Shape Model: Data From The Osteoarthritis Initiative.” Osteoarthritis and cartilage vol. 31, 1 (2023): 78–79. doi: 10.1016/j.joca.2023.01.020.

[11] Bowes, Michael A et al. “Machine-learning, MRI bone shape and important clinical outcomes in osteoarthritis: data from the Osteoarthritis Initiative.” Annals of the rheumatic diseases vol. 80,4 (2021): 502–508. Doi: 10.1136/annrheumdis-2020-217160.

[12] Kogan, Feliks et al. “PET/MRI of metabolic activity in osteoarthritis: A feasibility study.” Journal of magnetic resonance imaging : JMRI vol. 45,6 (2017): 1736–1745. doi:10.1002/jmri.25529.

[13] Goyal, Ananya et al. “Metabolic bone imaging and its relationship with biomechanics”. Osteoarthritis Imaging vol. 4,3 (2024). doi: 10.1016/j.ostima.2024.100242.

[14] Eckstein, Felix et al. “Detection of Differences in Longitudinal Cartilage Thickness Loss Using a Deep-Learning Automated Segmentation Algorithm: Data From the Foundation for the National Institutes of Health Biomarkers Study of the Osteoarthritis Initiative.” Arthritis care & research vol. 74,6 (2022): 929–936. doi:10.1002/acr.24539

[15] Eagle, Sonja et al. “Morphologic and quantitative magnetic resonance imaging of knee articular cartilage for the assessment of post-traumatic osteoarthritis.” Journal of orthopaedic research : official publication of the Orthopaedic Research Society vol. 35,3 (2017): 412–423. doi:10.1002/jor.23345

[16] Urish, Kenneth L et al. “Registration of Magnetic Resonance Image Series for Knee Articular Cartilage Analysis: Data from the Osteoarthritis Initiative.” Cartilage vol. 4,1 (2013): 20–27. doi:10.1177/1947603512451745

[17] Desai A, Barbieri M, Mazzoli V, Rubin E, Black M, Watkins L, Gold G, Hargreaves B, Chaudhari A. DOSMA: A deep-learning, open-source framework for musculoskeletal MRI analysis (Version v0.0.9, prerelease). Zenodo. Feb 2019. 10.5281/zenodo.2559549.

[18] Gatti, Anthony A et al. “ShapeMed-Knee: A Dataset and Neural Shape Model Benchmark for Modeling 3D Femurs.” medRxiv : the preprint server for health sciences 2024.05.06.24306965. 22 Oct. 2024, doi:10.1101/2024.05.06.24306965. Preprint.

[19] Gatti, Anthony A, KneePipeline (2025), GitHub Repository, https://github.com/gattia/KneePipeline.

[20] Sveinsson, B et al. “A simple analytic method for estimating T2 in the knee from DESS.” Magnetic resonance imaging vol. 38 (2017): 63–70. doi:10.1016/j.mri.2016.12.018

[21] Peterfy, C G et al. “The osteoarthritis initiative: report on the design rationale for the magnetic resonance imaging protocol for the knee.” Osteoarthritis and cartilage vol. 16,12 (2008): 1433–41. doi:10.1016/j.joca.2008.06.016

[22] Gatti, A.A., Maly, M.R. Automatic knee cartilage and bone segmentation using multi-stage convolutional neural networks: data from the osteoarthritis initiative. Magn Reson Mater Phy 34, 859–875 (2021). 10.1007/s10334-021-00934-z

[23] Peterfy, C G et al. “The osteoarthritis initiative: report on the design rationale for the magnetic resonance imaging protocol for the knee.” Osteoarthritis and cartilage vol. 16,12 (2008): 1433–41. doi:10.1016/j.joca.2008.06.016

[24] Desai AD, Schmidt AM, Rubin EB, et al. SKM-TEA: A Dataset for Accelerated MRI Reconstruction with Dense Image Labels for Quantitative Clinical Evaluation. Published online March 13, 2022. doi:10.48550/arXiv.2203.06823

[25] Kingma, D.P. and Ba, J. (2017) Adam: A Method for Stochastic Optimization. 1412.6980v9.

[26] Chollet, Francois, Keras (2015), https://keras.io.

[27] Gatti, Anthony A, pyMSKT (2023), GitHub Repository, https://github.com/gattia/pymskt.

[28] Eckstein, Felix, and Wolfgang Wirth. “Quantitative cartilage imaging in knee osteoarthritis.” Arthritis vol. 2011 (2011): 475684. doi:10.1155/2011/475684

[29] Gatti, Anthony A, ShapeMedKnee (2024), GitHub Repository, https://github.com/gattia/shapemedknee.

[30] Ansel, J., et al. “PyTorch 2: Faster Machine Learning Through Dynamic Python Bytecode Transformation and Graph Compilation.” Proceedings of the 29th ACM International Conference on Architectural Support for Programming Languages and Operating Systems, vol. 2, ACM, 2024, pp. 929–947. ISBN 9798400703850, 10.1145/3620665.3640366.

[31] Hawkins, R A et al. “Evaluation of the skeletal kinetics of fluorine-18-fluoride ion with PET.” Journal of nuclear medicine : official publication, Society of Nuclear Medicine vol. 33,5 (1992): 633–42.

[32] Harris, C.R., Millman, K.J., van der Walt, S.J. et al. Array programming with NumPy. Nature 585, 357–362 (2020). DOI: 10.1038/s41586-020-2649-2

[33] R. Beare, B. C. Lowekamp, Z. Yaniv, “Image Segmentation, Registration and Characterization in R with SimpleITK”, J Stat Softw, 86(8), doi: 10.18637/jss.v086.i08, 2018

[34] Ambellan, Felix et al. “Automated segmentation of knee bone and cartilage combining statistical shape knowledge and convolutional neural networks: Data from the Osteoarthritis Initiative.” Medical image analysis vol. 52 (2019): 109–118. doi:10.1016/j.media.2018.11.009

[35] Kessler, Dimitri A et al. “Segmentation of knee MRI data with convolutional neural networks for semi-automated three-dimensional surface-based analysis of cartilage morphology and composition”. Osteoarthritis Imaging vol. 2,2 (2022). doi: 10.1016/j.ostima.2022.100010

[36] Schmidt, Andrew M et al. “Generalizability of Deep Learning Segmentation Algorithms for Automated Assessment of Cartilage Morphology and MRI Relaxometry.” Journal of magnetic resonance imaging : JMRI vol. 57,4 (2023): 1029–1039. doi:10.1002/jmri.28365

[37] Tack, A et al. “Knee menisci segmentation using convolutional neural networks: data from the Osteoarthritis Initiative.” Osteoarthritis and cartilage vol. 26,5 (2018): 680–688. doi:10.1016/j.joca.2018.02.907

[38] Gaj, Sibaji et al. “Automated cartilage and meniscus segmentation of knee MRI with conditional generative adversarial networks.” Magnetic resonance in medicine vol. 84,1 (2020): 437–449. doi:10.1002/mrm.28111

[39] Norman, Berk et al. “Use of 2D U-Net Convolutional Neural Networks for Automated Cartilage and Meniscus Segmentation of Knee MR Imaging Data to Determine Relaxometry and Morphometry.” Radiology vol. 288,1 (2018): 177–185. doi:10.1148/radiol.2018172322

[40] Eckstein, Felix et al. “Clinical validation of fully automated laminar knee cartilage transverse relaxation time (T2) analysis in anterior cruciate ligament (ACL)-injured knees-on behalf of the osteoarthritis (OA)-Bio consortium.” Quantitative imaging in medicine and surgery vol. 14,7 (2024): 4319–4332. doi:10.21037/qims-24-194

[41] Wirth, Wolfgang et al. “Evaluation of an automated laminar cartilage T2 relaxation time analysis method in an early osteoarthritis model.” Skeletal radiology vol. 54,3 (2025): 571–584. doi:10.1007/s00256-024-04786-1

[42] Wirth, Wolfgang et al. “Accuracy and longitudinal reproducibility of quantitative femorotibial cartilage measures derived from automated U-Net-based segmentation of two different MRI contrasts: data from the osteoarthritis initiative healthy reference cohort.” Magma (New York, N.Y.) vol. 34,3 (2021): 337–354. doi:10.1007/s10334-020-00889-7

[43] Eckstein, Felix et al. “The design of a sample rapid magnetic resonance imaging (MRI) acquisition protocol supporting assessment of multiple articular tissues and pathologies in knee osteoarthritis.” Osteoarthritis and cartilage open vol. 6,3 100505. 23 Jul. 2024, doi:10.1016/j.ocarto.2024.100505

[44] Kellgren J H and Lawrence J S. “Radiological assessment of osteo-arthrosis.” Annals of the rheumatic diseases vol. 16,4 (1957): 494–502. doi:10.1136/ard.16.4.494

[45] Williams AA, Koltsov JCB, Brett A, He J, Chu CR. Using 3D MRI Bone Shape to Predict Pre-Osteoarthritis of the Knee 2 Years After Anterior Cruciate Ligament Reconstruction. Am J Sports Med. 2023;51(14):3677–3686. doi:10.1177/03635465231207615

[46] Haddock, Bryan et al. “Kinetic [18F]-Fluoride of the Knee in Normal Volunteers.” Clinical nuclear medicine vol. 44,5 (2019): 377–385. doi:10.1097/RLU.0000000000002533

[47] Kogan, Feliks et al. “PET-MRI: The promise of multi-tissue imaging of early disease mechanisms in osteoarthritis.” Osteoarthritis and cartilage vol. 33,1 (2025): 5–8. doi:10.1016/j.joca.2024.10.011

[48] Kogan, Feliks et al. “Multimodal positron emission tomography (PET) imaging in non-oncologic musculoskeletal radiology.” Skeletal radiology vol. 53,9 (2024): 1833–1846. doi:10.1007/s00256-024-04640-4

