## Supplementary Text for "Automating Imaging Biomarker Analysis for Knee Osteoarthritis Using an Open-Source MRI-Based Deep Learning Pipeline"

**Section 1: PET Image Dataset and Processing**

*Image Dataset*

Scan participants were hand-injected via an intravenous catheter with 92.5 MBq [^18^F]NaF, and scanned in a 3T Signa PET-MR scanner. Dynamic PET LST data was acquired for both knees for 30 minutes. From the LST data, images for the arterial input function (AIF), time activity curve (TAC), PET angiography (PETA), and static end of scan, were reconstructed. The reconstructed PET images had resolution 1.3x1.3x2.78 mm. For calculation of the image-derived arterial input function (AIF), dynamic PET frame times of 2 × 5 seconds, 20 x 1 seconds, 10 × 10 seconds, 10 x 30 seconds, 5 x 1 minutes, and 9 x 2 minutes were reconstructed using time-of-flight–ordered subset expectation maximization with 3 iterations with corrections for decay, attenuation, scatter, random, and dead time. Time activity curves (TAC) of bone PET uptake were determined using dynamic PET time frames of 6 x 10 seconds, 10 x 1 minutes, and 9 x 2 minutes with the same corrections. For the PET angiography images, static PETA images were reconstructed 15 seconds after the bolus delivery. Similarly, static end of scan images were reconstructed from the last five minutes of the dynamic scan, 25-30 minutes after the bolus delivery.

*Image Processing Pipeline*

In-house MATLAB codes were used for pharmacokinetic analysis, which is shared through an open-source GitHub repository, dyNaFKnee (<https://github.com/ananya-goyal/dyNaFknee>). Descriptions of each code and their corresponding function is below:

Make_AIF.m: Dynamic 4D AIF images and 3D PETA images are used to segment blood vessels and extract arterial input function from segmented region. For segmentation, peak activity is identified, background noise is removed, and area under the curve and gradient analysis are computed to refine the vessel selection. Then, mean PET intensity values within the segmented vessel region are computed over time.

TAC_coreg.m: Perform co-registration of 4D TAC (Time-Activity Curve) images to a single reference frame. We automatically segment the two legs and register them separately using monomodal rigid registration.

Import_ROI.m: Load qDESS-based segmentations into the matrix.

TAC.m: Loads the pre-defined ROIs and co-registered PET TAC data. Uses the function makeTACgraph to calculate TAC curves for the specified ROIs.

ROI_seg.m: Loads AIF, TAC data, along with segmentations (ROIS) into the matrix. Divide ROIs into left and right regions for each scan.

ROI_disp_kin.m: Using COMKAT, for each ROI, compute TAC, and apply a kinetic model. Fits a two-tissue compartment model to the TAC using the AIF and calculates parameters: rate constants (K1, k2, k3), blood flow delay and tissue uptake (Kipat, DV, etc.).
